# High incidence of glucocorticoid-induced hyperglycaemia in inflammatory bowel disease; metabolic and clinical predictors identified by machine learning

**DOI:** 10.1101/2020.06.22.20137356

**Authors:** Martin McDonnell, Richard J Harris, Florina Borca, Tilly Mills, Louise Downey, Suranga Dharmasiri, Mayank Patel, Benjamin Zaré, Matt Stammers, Trevor R Smith, Richard Felwick, Fraser Cummings, Hang T.T. Phan, Markus Gwiggner

## Abstract

**Background:** Glucocorticosteroids (GC) are long-established, widely used agents for induction of remission in inflammatory bowel disease (IBD). Hyperglycaemia is a known complication of GC treatment with implications for morbidity and mortality. Published data on prevalence and risk factors for GC-induced hyperglycaemia in the IBD population are limited. We prospectively characterise this complication in our cohort, employing machine-learning methods to identify key predictors of risk.

**Methods:** We conducted a prospective observational study of IBD patients receiving intravenous hydrocortisone (IVH). Electronically triggered three times daily capillary blood glucose (CBG) monitoring was recorded alongside diabetes mellitus (DM) history, IBD biomarkers, nutritional and IBD clinical activity scores. Hyperglycaemia was defined as CBG ≥11·1mmol/L and undiagnosed DM as HbA1c ≥48 mmol/mol. Random Forest regression models were used to extract predictor-patterns present within the dataset.

**Findings:** 94 consecutive IBD patients treated with IVH were included. 60% (56/94) of the cohort recorded an episode of hyperglycaemia, including 57% (50/88) of those with no prior history of DM, of which 19% (17/88) and 5% (4/88) recorded a CBG ≥14mmol/L and ≥20mmol/L, respectively. The Random Forest models identified increased CRP followed by a longer IBD duration as leading risk predictors for significant hyperglycaemia.

**Interpretation:** Hyperglycaemia is common in IBD patients treated with intravenous GC, therefore CBG monitoring should be included in routine clinical practice. Machine learning methods can identify key risk factors for clinical complications. Physicians should consider steroid-sparing strategies in high-risk patients such as those with high admission CRP or a longer IBD duration. There is an emergent case for research to explore steroid-free treatment regimens for hospitalised patients with severe IBD flares.

**Evidence before this study:** Glucocorticosteroids (GC) are long-established induction agents in the management of inflammatory bowel disease (IBD). They are recommended first-line therapy in consensus guidelines and prescribing remains widespread, with an estimated 30% of IBD patients exposed annually. Hyperglycaemia is a known complication of GC and has been linked to increased length of hospital stay, morbidity and mortality. Small case series of GC treated medical patients suggest a higher risk of hyperglycaemia in the hospitalised population but have suffered from a lack of systematic blood glucose monitoring.

**Added value of this study:** This is the first study utilising prospective, systematic monitoring of capillary blood glucose (CBG) to determine the frequency of hyperglycaemia in a GC-treated hospitalised IBD population. We report that more than half of IBD patients without prior diabetes mellitus treated with intravenous hydrocortisone (IVH), will develop hyperglycaemia (CBG ≥11·1mmol/L). Random Forest regressors pinpointed CRP and IBD duration as the strongest predictor of this adverse outcome.

**Implications of all the available evidence:** Hyperglycaemia is a common complication of IVH therapy in hospitalised IBD patients, particularly in those with high inflammatory burden. The monitoring and management of this complication, which has potential implications for the morbidity, mortality and subsequent risk of diabetes diagnosis should become part of routine clinical practice.

## INTRODUCTION

In the 1950’s Truelove and Witts performed the first randomised controlled trial of glucocorticosteroids (GC) in active ulcerative colitis (UC), demonstrating a reduction in mortality from 10·9% to 4·6% after two months of treatment.^1^ 70 years later, GC remain a common treatment for inflammatory bowel disease (IBD); a recent UK multi-centre study identified 30% of patients were exposed to GC in the preceding 12 months.^2^ GC are first-line induction agents for moderate to severe Crohn’s Disease (CD) and UC in consensus guidelines and intravenous GC are widely used as the initial medical therapy for IBD patients admitted to hospital with severe disease flares.^3,4^

The mechanism of action of GC is mediated via the cytosolic glucocorticoid receptor (cGR). After GC binding, cGR-associated proteins dissociate, and the GC-cGR complex translocates to the nucleus. The nuclear GC-cGR complex binds to DNA, upregulating the transcription of anti-inflammatory cytokines (e.g. IL-10, annexin 1) and downregulating transcription of pro-inflammatory mediators (e.g. IL-1α, IL-1β, IL-8, TNFα). More rapid, non-genomic, effects occur via both the cGR dissociated proteins and likely via other novel transmembrane receptors.^5^ Steroid-induced hyperglycaemia is mediated by decreased β-cell insulin production, increased insulin resistance and increased gluconeogenesis.^6^

Studies in other GC-treated populations demonstrate both short and long-term severe adverse effects including ocular, musculoskeletal, cardiovascular, metabolic and infectious complications.^7^ In addition, de-novo hyperglycaemia and deterioration of glycaemic control in those with pre-existing diabetes mellitus (DM) have been recognised adverse effects of GC since the 1950s.^8^ The specific risk in IBD hospitalised patients with severe IBD flares, however, is not known.

Hyperglycaemia has been associated with increased length of stay, morbidity and mortality in non-critical care medical and surgical patients.^9^ At least 67 cases of GC induced metabolic emergencies including ketoacidosis and hyperosmolar hyperglycaemic states have been reported, 11 of which were fatal.^10^ Following 2 cases of GC-related diabetic crisis at our institution in IBD patients with no prior diabetes, electronically triggered systematic capillary blood glucose (CBG) monitoring was introduced for all prescribed intravenous hydrocortisone (IVH). This allowed the systematic study of hyperglycaemia in hospitalised IBD patients receiving IVH.

## MATERIALS AND METHODS

We conducted a prospective observational study of consecutive admissions of IBD patients treated with IVH between October 2017 and December 2018. Included subjects had a confirmed diagnosis of IBD (either historic or during admission episode) and received at least four doses of 100mg IVH over 24 hours.

Subjects had three times daily CBG monitoring automatically triggered by electronic prescription of IVH. CBG results were automatically digitally captured in the electronic patient record. Predictor variables included admission IBD severity score (Harvey Bradshaw Index or partial Mayo Score), serum biochemistry, HbA1c, faecal calprotectin, C-reactive protein, potential DM risk factors (BMI, family history, concomitant medications), self-reported weight loss and MUST (Malnutrition Universal Screening Tool) score.^11^

Patients developing a CBG ≥14mmol/l were referred to the diabetes team for consideration of treatment. Undiagnosed DM was excluded by baseline HbA1c <48 mmol/mol or a pre-IVH CBG <11·1mmol/L.^12^ Biochemical follow up was performed at one year to assess repeat HbA1c measurements and serum and faecal biomarker assessments.

Laboratory values outside the quantification limit were substituted with the upper/lower limit value. Categorical and continuous predictors were handled and formatted appropriately. Data were analysed using Python v3·7·4 and R v3·6·1. Descriptive statistics were applied to cohorts, and Pearson correlations between independent and dependent variables were ascertained with matplotlib/seaborn heat-maps. Random Forest (RF) machine learning models using the scikit-learn package v0·22·1 were constructed to regress the phenotypic and admission clinical predictors.^13,14^ Pairwise correlations between input features (see supplementary data) were identified to eliminate similar features. Then multivariable RF models were constructed and optimised using random grid search. Subsequently 5-fold cross-validation with a train-test split was used to test effectiveness at predicting glucose levels in the internal holdout set. Mean squared error was the reported metric.

The protocol was reviewed and approved by the UK Health Research Authority - reference 19/HRA/0033.

## RESULTS

### Demographics

94 IBD inpatient episodes met the inclusion criteria; 54 for CD, 36 for UC and 4 for IBD-as yet unclassified (IBDU). The median length of IVH treatment was four days (range 1-11).

5/94 episodes were for patients with a prior diagnosis of Type 2 DM. One further subject had a pre-GC treatment CBG of 13·9mmol/L and was thus reclassified as undiagnosed Type 2 DM.

**Table 1:** Demographics.

|  |  | **CD** | **IBDU** | **UC** |
| --- | --- | --- | --- | --- |
|  | **n** | **54** | **4** | **36** |
| **Age** |  | 42.2 (37.9 - 46.5) | 49.8 (28.4 - 71.1) | 43.1 (37.0 - 49.2) |
| **Gender** | **Female** | 25 (46.3) | 2 (50.0) | 19 (52.8) |
| **BMI** | **<18.5** | 6 (11.1) | 1 (25.0) | 3 (8.3) |
|  | **18.5 - 25** | 28 (51.9) | 2 (50) | 16 (44.4) |
|  | **25-30** | 13 (24.1) | 1 (25) | 14 (38.9) |
|  | **>30** | 7 (13.0) | 0 (0.0) | 3 (8.3) |
| **MUST Score** | **0** | 17 (31.5) | 1 (25.0) | 17 (48.6) |
|  | **1** | 13 (24.1) | 1 (25.0) | 10 (28.6) |
|  | **2** | 13 (24.1) | 2 (50.0) | 4 (11.4) |
|  | **≥3** | 11 (20.4) | 0 (0.0) | 4 (11.4) |
| **Length of diagnosis (years)** |  | 7.3 (4.6 - 10.0) | 3.3 (0 - 8.4) | 5.0 (3.2 - 6.8) |
| **History of Diabetes** | **No** | 49 (90.7) | 4 (100.0) | 35 (97.2) |
|  | **Type 2 DM** | 5 (9.3) | 0 (0.0) | 1 (2.8) |
| **Admission HBA1C** |  | 38.5 (35.5 - 41.5) | 37.0 (33.1 - 40.8) | 37.7 (35.8 - 39.6) |
| **Admission Calprotectin** |  | 2854 (1980 - 2298) | 2491 (NA) | 3159 (2302 - 4071) |
| **Admission CRP** |  | 75.9 (50.5 - 101.3) | 138.0 (106.7 - 169.3) | 88.9 (53.6 - 124.1) |
| **PMS** |  | .. | 5.50 (4.5 - 6.5) | 7.78 (7.2 - 8.3) |
| **HBI** |  | 14.7 (13.0-16.5) | .. | .. |
| **Current Treatment** | **Oral steroids** | 8 (14.8) | 1 (25) | 6 (16.7) |
|  | **Oral 5ASA** | 2 (3.7) | 2 (50) | 20 (55.6) |
|  | **Immunomodulator** | 18 (33.3) | 0 (0) | 6 (16.7) |
|  | **Anti-TNF** | 10 (19.2) | 0 (0) | 10 (29.4) |
|  | **Vedolizumab** | 2 (3.8) | 0 (0) | 0 (0.0) |
|  | **Ustekinumab** | 7 (13.5) | 0 (0) | 0 (0.0) |
| **Smoking** | **Current** | 12 (23.5) | 0 (0) | 0 (0.0) |
|  | **Ex-smoker** | 9 (17.7) | 1 (25.0) | 13 (36.1) |
|  | **Never smoked** | 30 (58.8) | 3 (75.0) | 23 (63.9) |

For categorical variables n (%) shown, for continuous variables mean (95% Confidence Interval) shown. Abbreviations: '..' = not applicable, MUST (Malnutrition Universal Screening Tool), BMI (Body Mass Index), PMS (Partial Mayo Score), HBI (Harvey Bradshaw Index), 5-ASA (5-aminosalicylic acid), Anti-TNF (anti-Tumour Necrosis Factor), Immunomodulator (azathioprine, mercaptopurine or methotrexate).

### Incidence and management of hyperglycaemia

The overall global incidence of hyperglycaemia in our cohort of hospitalised IBD patients receiving IVH was 60% (56/94), with intervention for diabetes initiated in 20% (11/56) of these patients. For those without a prior diagnosis of DM, 57% (50/88) developed diabetic-range CBG (≥11·1mmol/L) while 19% (17/88) and 5% (4/88) developed a CBG ≥14mmol/L and ≥20mmol/L, respectively. 25/50 of those with de-novo hyperglycaemia had CD, 22/50 UC, and 3/50 IBDU with a combined mean age of 44·8 years (Standard Deviation 17·4). In patients without a prior diagnosis of DM 14% (7/50) had diabetic interventions; 6/50 started an oral hypoglycaemic and 1/50 commenced insulin therapy.

All six patients with prior DM (5 CD and 1 UC) had an episode of hyperglycaemia and 4/6 recorded a CBG ≥20mmol/L. 50% (3/6) were started on insulin, and 17% (1/6) commenced oral hypoglycaemic therapy.

Of the 50 admissions in patients without prior DM recording a CBG ≥11·1mmol/L, 22% (11/50) were preceded directly by a course of oral steroids compared to 8% (3/38) without hyperglycaemia (p = 0·09, Fisher’s exact test). In 60% (30/50) peak CBG occurred on day two of IVH treatment, but in 14% (7/50) of cases peak CBG was not reached until seven days after the first dose.

During index admission, 54% (30/56) of hyperglycaemic patients and 50% (19/38) of normoglycaemic patients were started on biologic therapy. Four patients underwent emergency inpatient colectomy, all of whom had recorded a hyperglycaemic episode. Mean (± 95% Confidence Interval) length of stay was 12·6 (± 3·0) days for hyperglycaemic patients and 9·1 (± 2·0) days for normoglycaemic patients (p = 0·053, t-test).

Physicians’ decision making was influenced by steroid-induced hyperglycaemia, particularly in those with a persistent CBG ≥14mmol/L (trigger for diabetes specialist review). Conventional post-IVH eight-week tapering course of oral prednisolone prescribing differed. Only 30% of patients with CBG ≥14mmol/L received oral prednisolone without intervention (e.g. diabetic medication or rapid steroid taper) versus 58% of those with normoglycemia.

### Predictors of hyperglycaemia

Maximum CBG was positively correlated with CRP, platelet count, and HbA1c and negatively correlated with haemoglobin (see correlation heat-map in supplementary data). For non-diabetic patients, the RF model was fit 500 times with a maximum depth of 90 trees using a random grid search method. CRP was the most critical predictor identified, followed by disease duration, platelet count, admission haemoglobin and HbA1C. The final mean squared error of the optimised non-diabetic patient model was 1·876.

The variables and their respective importance of contribution to the RF modelling are shown in figure 2.

**Figure 1:**
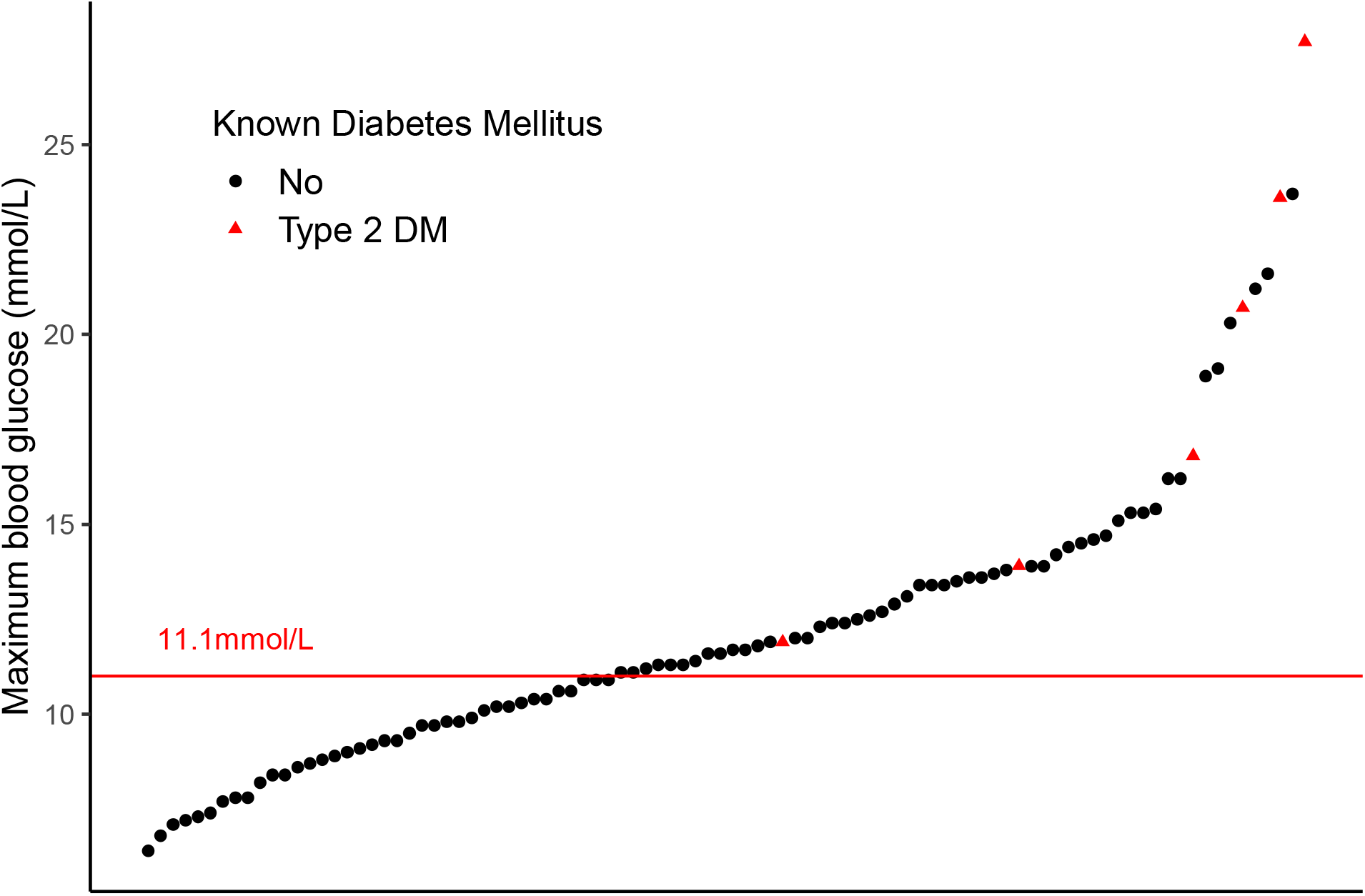
Maximum recorded CBG for each admission plotted in ascending order.

**Figure 2:**
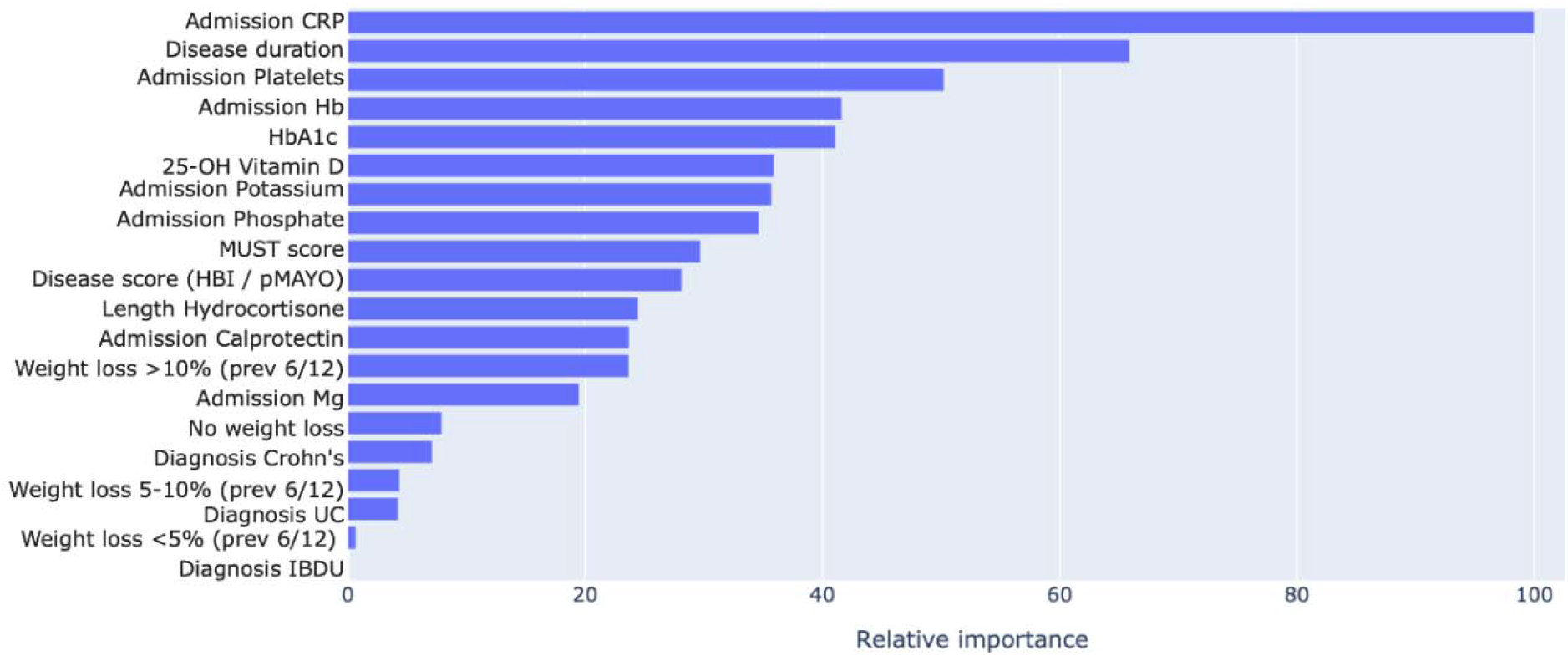
Relative variable importance for Random Forest modelling of maximum CBG (subjects with history of DM excluded)

RF models optimised by grid search will rank the relative importance of each variable against the most critical variable (in this case, CRP). This provides a sense of how much each variable contributes to the predictive power of the overall model. In this instance, the length of IVH usage and metabolic variables are not as crucial to predicting hyperglycaemia as blood markers of inflammation.

Once the optimised RF model had been developed, blood glucose predictions were made on the holdout set using 5-fold cross validation to reduce overfitting.

Figure 3 reveals that the test-set predictions were comparable to the training set predictions. This pattern was present across all folds of the cross-validation; this graph is given only as an example of the model’s performance.

**Figure 3:**
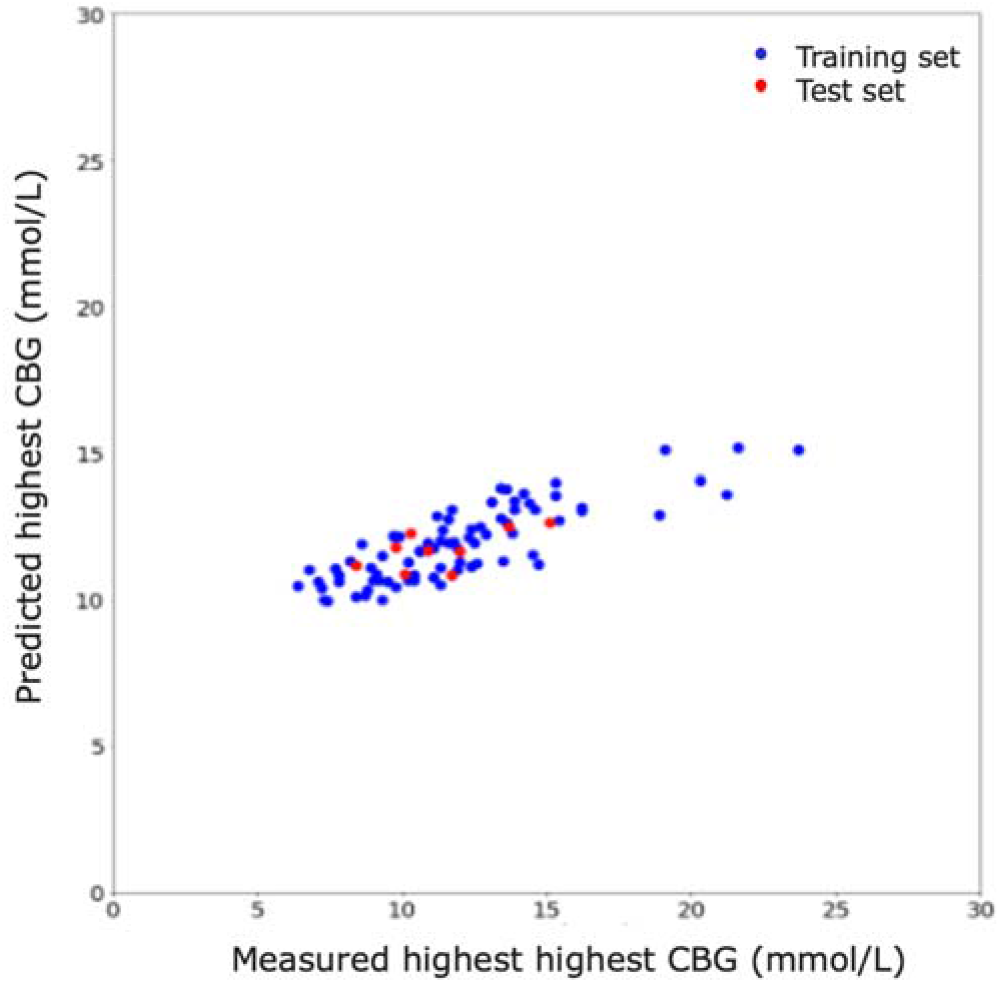
Predicted highest CBG vs measured highest CBG using model (subjects with history of DM excluded)

When the RFR model included the six patients with DM, the leading predictor of CBG elevation became HbA1c. Due to high CBG elevation (≥20mmol/L) in subjects with known diabetes, particularly those with evidence of inadequate control by admission HbA1c, the RFR modelling shown above excluded these patients. (See supplementary data) for the variable importance, correlation matrix and model with the patients with previous diabetes included. Their inclusion increased the mean squared error to 2·158, which is not surprising given the differing pre-admission characteristics in this sub-population.

### Additional reported side effects and metabolic instability

32% (30/94) of episodes included a patient-reported symptom of anxiety or mood change, 11% (10/94) of poor sleep and 10% (9/94) a report of appetite change. Electrolyte depletion, like hyperglycaemia, is a marker of metabolic and nutritional dysregulation and was common in the whole cohort with no statistically significant differences between hyperglycaemic and normoglycemic patients. 62% (58/94) recorded a serum potassium below the lower limit of our laboratory reference range (<3·5mmol/L), 71% (67/94) low serum magnesium (<0·7mmol/l) and 84% (79/94) low serum phosphate (<0·8mmol/L). See supplementary data.

66% (37/56) of hyperglycaemic patients and 57% (21/38) of normoglycaemic patients reported weight loss in the six months before admission. All patients were routinely assessed by a specialist dietitian who prescribed nutritional support as indicated; 53/94 had oral nutritional supplements, 5/94 nasogastric feeding and 5/94 parenteral nutrition. There was no significant difference in the proportion of hyperglycaemic versus normoglycaemic patients receiving nutritional therapy.

### One-year follow-up

There was a significant decrease in faecal calprotectin over the 12-month follow-up. For those without hyperglycaemia during the acute episode, there was a trend to lower mean calprotectin at 12 months, 210 (± 707) versus 738 (± 640 p=0·18 t-test). In non-diabetic patients with raised calprotectin at admission and paired results 35% (6/17) normalised by 12 months in those who had hyperglycaemia and 75% (9/12) normalised in those without hyperglycaemia p=0·06 (Fisher’s exact test). There was no significant change in HbA1c over the one-year follow-up for any groups.

## DISCUSSION

Our data demonstrate that hyperglycaemia is common in IBD patients treated with IVH. By instituting electronically triggered regular CBG monitoring of IBD patients treated with IVH, we have uncovered the high prevalence of this relevant complication. Our monitoring regime identified patients who required diabetic intervention or modification of their treatment in response to hyperglycaemia. The void of data regarding steroid induced diabetes in IBD suggests that in current practice it may often be under-recognised, and thus under-treated, potentially exposing patients to the risk of complications such as increased length of stay and infection. Hyperglycaemia has also been mechanistically linked to intestinal barrier dysfunction in both murine models and non-IBD human subjects.^15^ Through this mechanism, it is at least theoretically possible to link hyperglycaemia directly to intestinal inflammation and IBD outcomes.^16^

Until now there has been no accurate prospective evidence of the extent and severity of IVH-induced hyperglycaemia in IBD. A previous retrospective study of steroid side effects in elderly Crohn’s disease patients estimated an incidence of just 17%, but did not have systematic CBG monitoring.^17^ Likewise, studies of GC-treated rheumatology patients report the occurrence of hyperglycaemia at 9 - 42%.^18-20^ A potential explanation for the higher frequency seen here is that our cohort only included hospitalised patients, who were both monitored frequently and likely to have had more significant systemic inflammation. The incidence we report is similar to that in a smaller study of a more elderly, heterogeneous group of hospitalised patients, prescribed GC.^21^

Machine learning techniques can be applied to clinical problems for risk prediction. We chose RF as it is a powerful ensemble machine learning method for predictive modelling, known to perform better than other methods on smaller datasets. Random Forests work by constructing a large number of smaller decision trees and averaging the output of individual trees and are capable of capturing complex dependency patterns among multi-variate input features.

The RF model accurately predicted the degree of hyperglycaemia for those without a history of DM. Principal independent determinants were admission CRP, length of disease followed by thrombocytosis and anaemia, suggesting that systemic inflammatory burden and duration of IBD determines the risk of clinically significant hyperglycaemia in IVH-treated IBD patients. No data of historic GC prescriptions were available so we cannot speculate further on whether the relationship with disease duration is a consequence of past GC exposure or a consequence of the disease process itself.

By using RF for early identification of those most at risk of significant GC-induced hyperglycaemia, alternative treatment strategies can be considered, including a rapid steroid taper, early use of biologics and avoidance of oral steroids in those demonstrating adequate response to biologic induction. Awareness of significant steroid-induced diabetes in this cohort appeared to influence physicians’ decisions with an observed reduction in follow-on prescriptions for conventional steroid taper.

Although hyperglycaemia appears to resolve on stopping GC, this is not always the case. A large population-based study associated previous oral GC with an increased incidence of future DM and in rheumatic diseases cumulative GC dose has been shown to be a risk factor for DM.^22,23^ The IBD population, regardless of GC exposure, may be at higher risk of DM. A recent Danish population-wide cohort study suggests a higher incidence of type 1 diabetes amongst subjects with IBD and a Korean cohort study found IBD patients under 40 to have an increased rate of incident diabetes diagnosis over five years, compared to age, sex and BMI-matched controls.^24,25^

This study has limitations. While we note that hyperglycaemia occurred during the admission of subjects treated with IVH, we do not know whether another aspect of the disease or its treatments was the principal cause. It is known that de-novo hyperglycaemia may also occur in hospitalised patients independently of GC as a result of the neurohormonal milieu of acute illness.^26^ As current guidelines recommend the use of high dose GC in moderate to severe acute IBD, there was no comparable cohort of patients managed without IVH treatment.^3^ Despite an attempt to measure baseline HbA1c in all patients starting steroids, 29% did not have this checked, and 3% had neither HbA1c nor CBG pre-IVH, so some cases of undiagnosed diabetes mellitus may have been missed. Future research should aim to validate the model in a wider population and determine any effect of hyperglycaemia on clinical outcomes, which our study was not powered to achieve.

Intravenous GC have undoubtedly saved the lives of many IBD patients. However, there is significant associated morbidity, as acknowledged seven decades earlier by Truelove and Witts in their seminal work.^1^ It is questionable if GCs would be licensed in the regulatory framework currently applied to newer agents, given their side effect profile. In the ever-evolving IBD therapeutic landscape, it may be necessary to re-evaluate the position of GCs in the management of IBD. In UC, ciclosporin and infliximab are of equivalent efficacy as ‘rescue’ therapy for those demonstrating inadequate response to IVH; whether and for how long the preceding and concomitant GC course should be given for is not known.^27^ The JAK inhibitor tofacitinib has demonstrated onset within three days in moderate to severe UC and has been used effectively alongside shorter IVH courses and budesonide as induction therapy in an acute severe UC case series.^28,29^

In acute CD the evidence for the benefit of GC to induce remission is less secure.^4^ Nutritional therapies can if given as exclusive enteral nutrition, be of similar efficacy as CD induction agents to oral GC. They may offer an alternative treatment strategy for those in whom steroids are contraindicated.^30^ When co-prescribed for a primary nutritional indication in our cohort, nutritional therapies did not increase the risk of hyperglycaemia.

Our data supports the inclusion of CBG monitoring into standard clinical practice for IBD patients receiving IVH. Machine learning identified patients with a high inflammatory disease burden and a longer disease duration as being at the greatest risk of significant hyperglycaemia. Given the common complications of GCs and an enlarging landscape of biological, small molecule and nutritional therapies, there is an emergent case for researching steroid-free treatment strategies for acute IBD.

## Data Availability

The data and algorithm is available on request from the author.

## ACKNOWLEDGEMENTS

We would like to thank the diabetes multidisciplinary specialist team at UHS for helping to look after these patients.

## Contribution statement

MM, RJH, SD and MG were responsible for the original concept and planning of the study. MM, RJH, TM, SD, BZ and LD were responsible for clinical data collection and analysis. FB and HP were responsible for data extraction, analysis and modelling. RJH and MM contributed equally to this work and drafted the manuscript, which RF, TS, MP, JRFC, HP, MS and MG critically reviewed and revised.

## SUPPLEMENTARY DATA

### Input features for RF model

MUST score, weight loss during the past six months (<5%, 5-10%, >10%), diagnosis (CD, UC, IBDU), time since diagnosis, admission faecal calprotectin and admission measurements of serum haemoglobin, platelets, magnesium, phosphate, potassium and CRP.

### GridSearch optimised parameters of the RF model

- For patients with no history of DM: ‘n_estimators’: 733, ‘min_samples_split’: 10, ‘min_samples_leaf’: 4, ‘max_features’: ‘sqrt’, ‘max_depth’: 90, ‘bootstrap’: True
- For all patients regardless of DM history: ‘n_estimators’: 3400, ‘min_samples_split’: 2, ‘min_samples_leaf’: 4, ‘max_features’: ‘sqrt’, ‘max_depth’: 80, ‘bootstrap’: False

**Supplementary figure 1:**
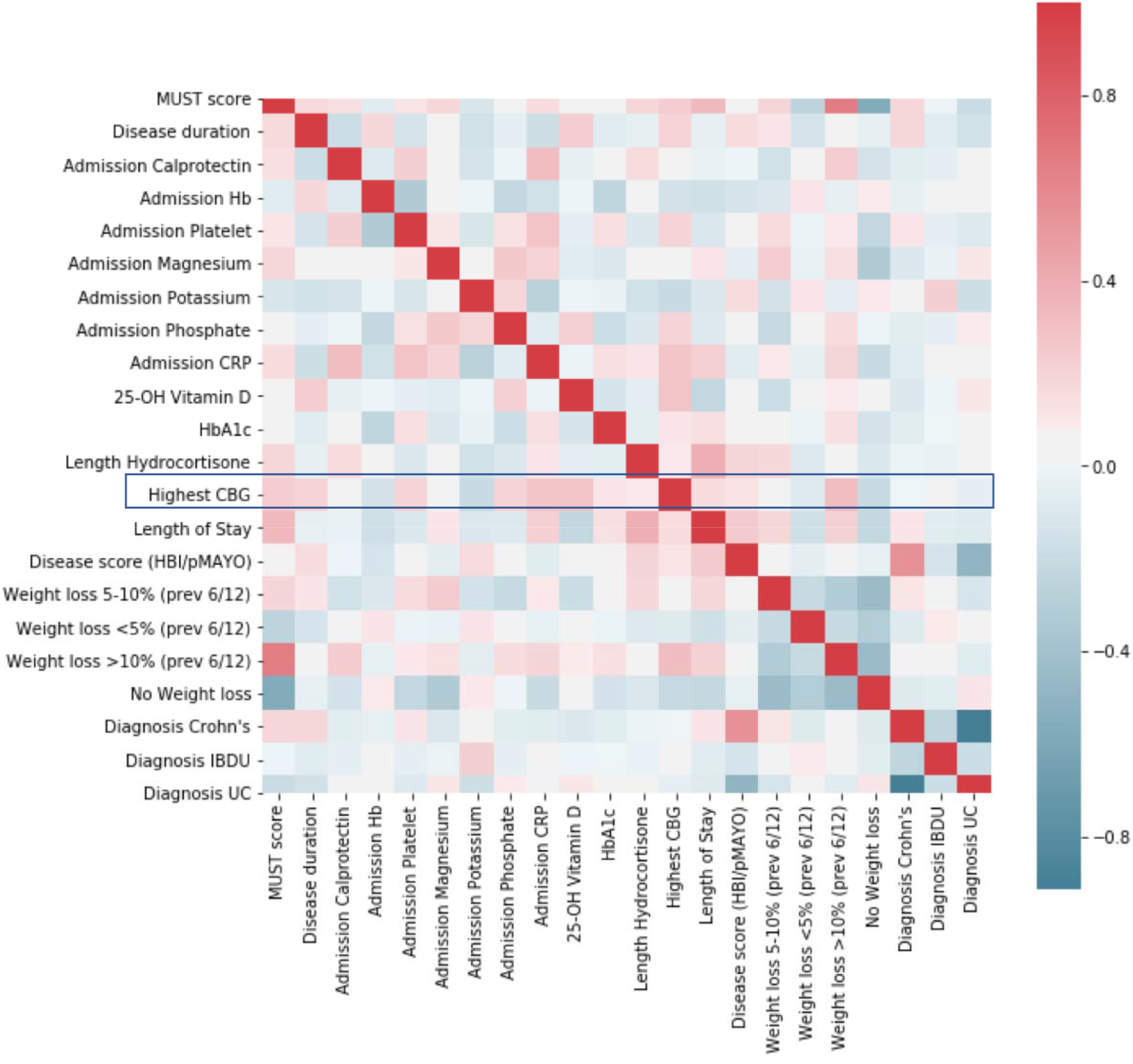
Correlation heat map of Random Forest parameters (subjects with history of DM excluded)

**Supplementary figure 2:**
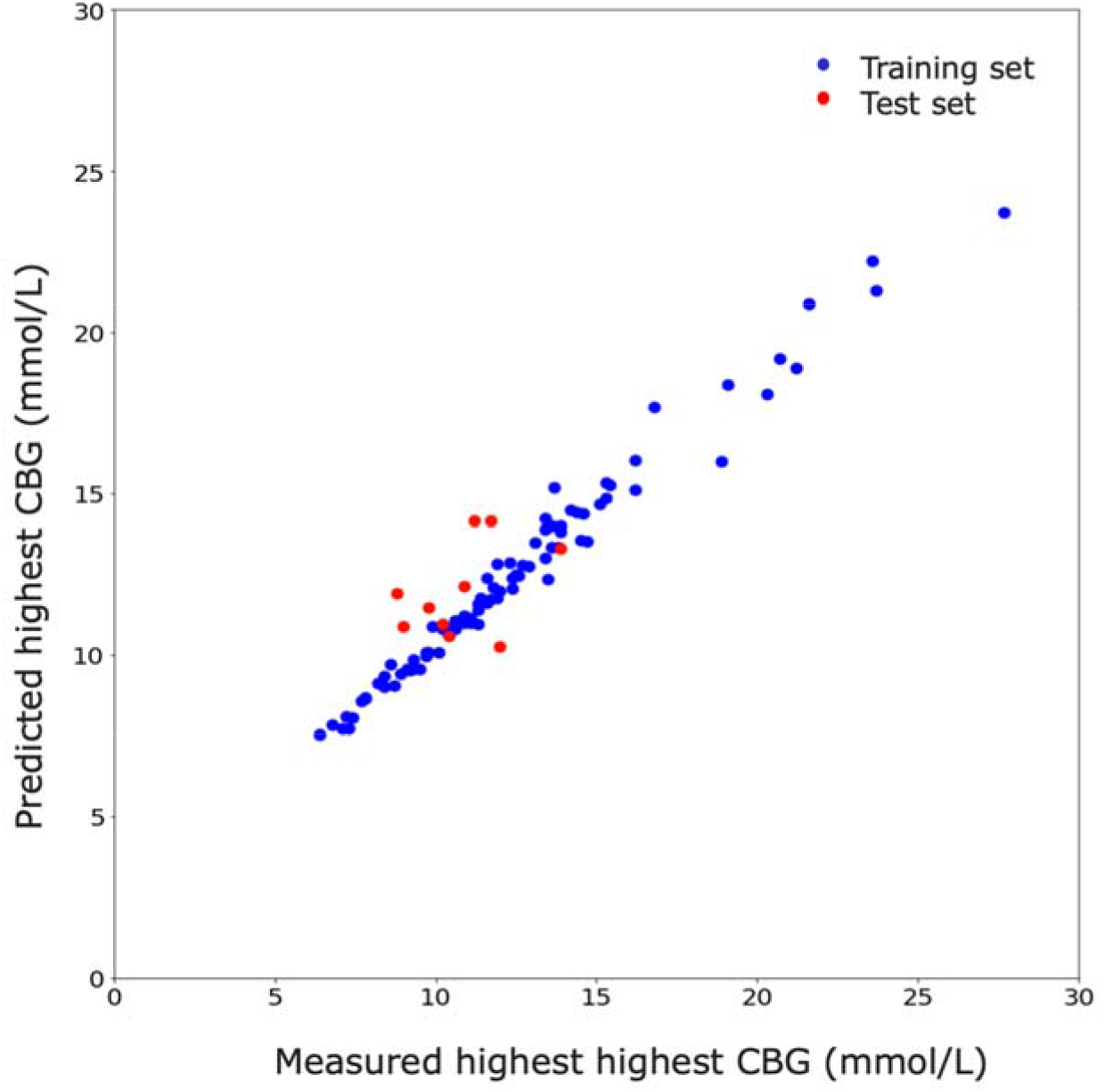
Predicted highest CBG vs measured highest CBG (all patients)

**Supplementary figure 3:**
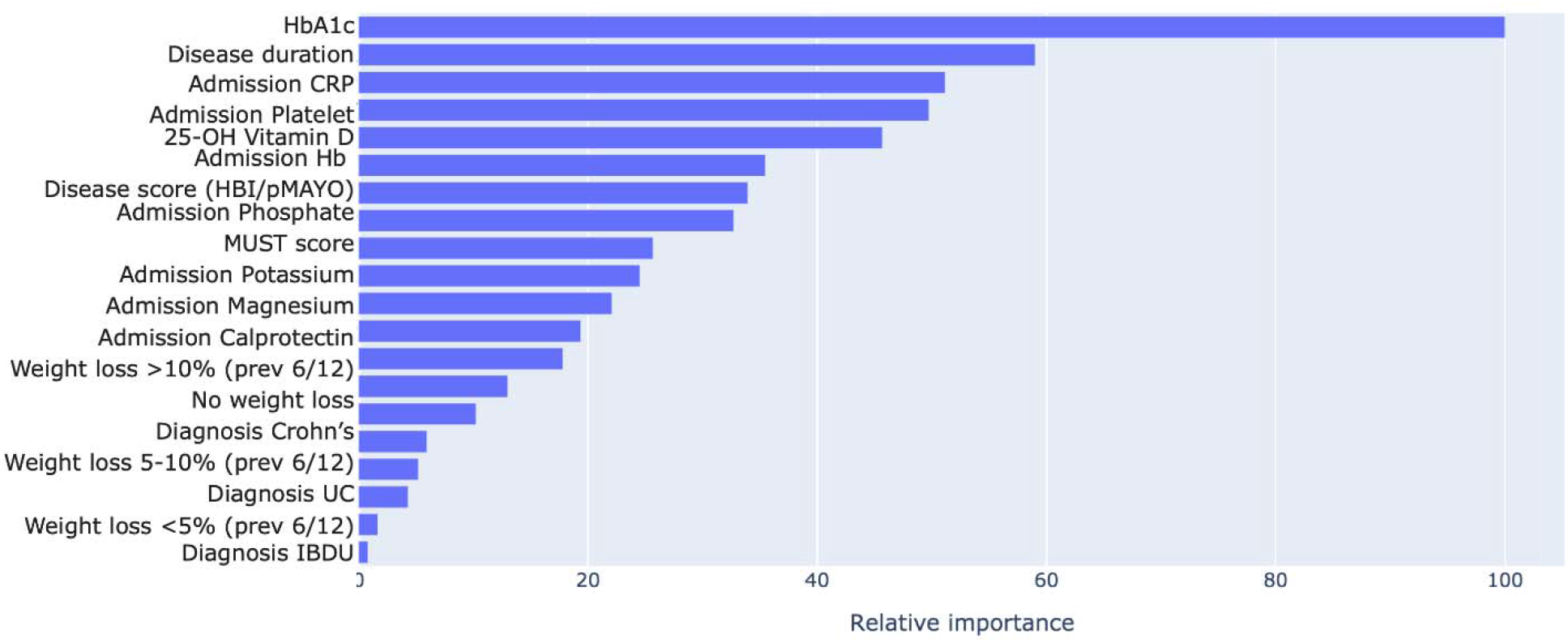
Variable importance for Random Forest modelling (all patients)

**Supplementary figure 4:**
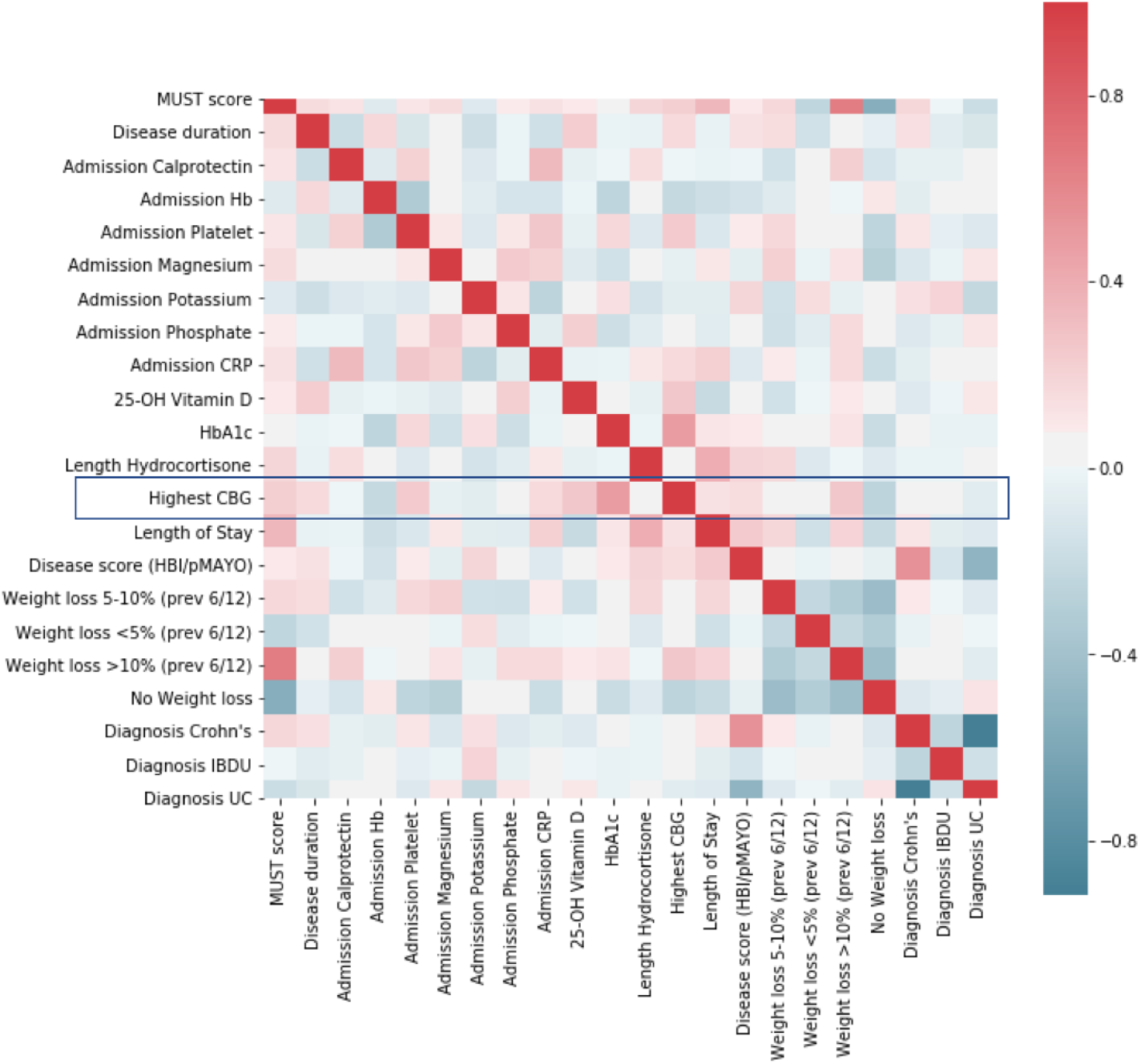
Correlation heat map of Random Forest parameters (all patients)

**Supplementary figure 5:**
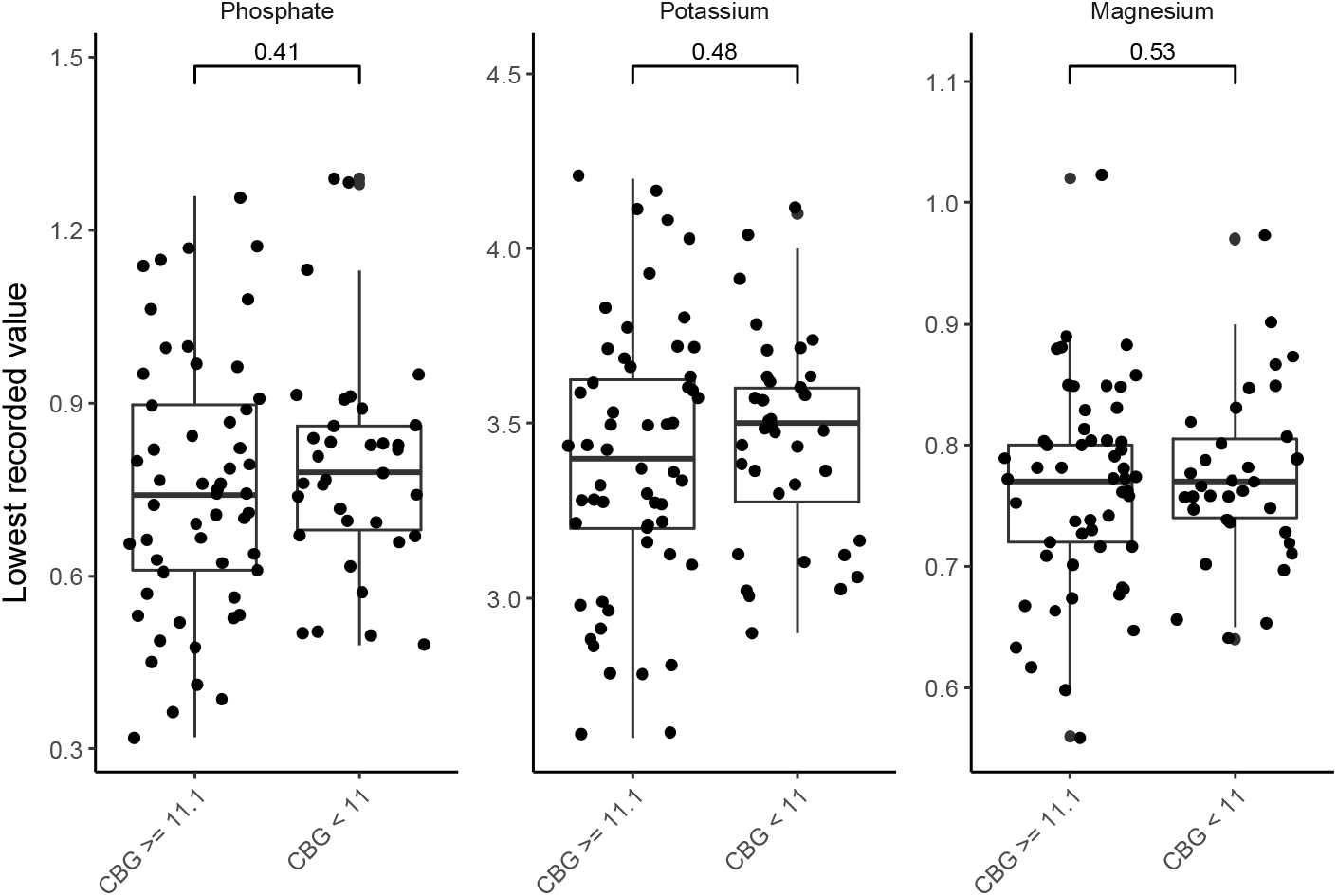
Lowest recorded electrolyte measurements (all patients) stratified by hyperglycaemia (maximum capillary blood glucose (CBG) ≥ 11mmol/L) or normoglycaemia (maximum CBG < 11mmol/L).

**Supplementary figure 6:**
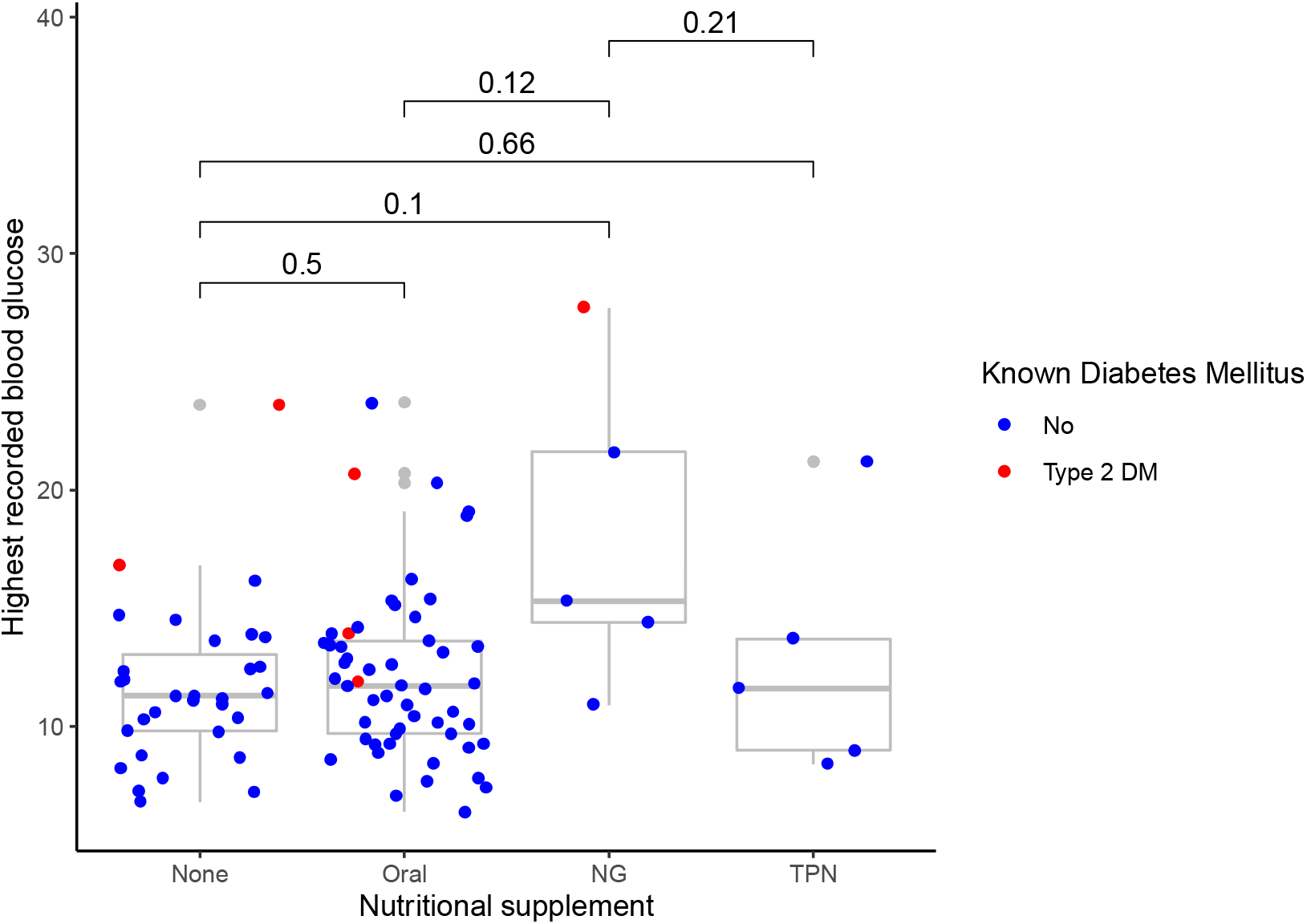
Highest recorded blood glucose stratified by nutritional supplements given (all patients) NG = Nasogastric tube feeding, Oral = Oral nutritional supplements, TPN = total parenteral nutrition

